# Graphene Multiplexed Sensor for Point-of-Need Viral Wastewater-Based Epidemiology

**DOI:** 10.1101/2024.03.18.24304492

**Authors:** Michael Geiwitz, Owen Rivers Page, Tio Marello, Narendra Kumar, Stephen Hummel, Vsevolod Belosevich, Qiong Ma, Tim van Opijnen, Bruce Batten, Michelle M. Meyer, Kenneth S. Burch

**Affiliations:** Department of Physics, Boston College, Chestnut Hill, MA 02467, USA; Department of Biology, Boston College, Chestnut Hill, MA 02467, USA; GRIP Molecular Technologies, Inc., 1000 Westgate Drive, Saint Paul, MN 55114, USA; Department of Chemistry and Life Science, United States Military Academy, West Point, NY 10996, USA

**Keywords:** SARS-CoV-2, Influenza, Respiratory syncytial virus, Graphene field effect transistor (GFET), Caffeine, Aptamer

## Abstract

Wastewater-based epidemiology (WBE) can help mitigate the spread of respiratory infections through the early detection of viruses, pathogens, and other biomarkers in human waste. The need for sample collection, shipping, and testing facilities drives up the cost of WBE and hinders its use for rapid detection and isolation in environments with small populations and in low-resource settings. Given the ubiquitousness and regular outbreaks of respiratory syncytial virus, SARS-CoV-2 and various influenza strains, there is a rising need for a low-cost and easy-to-use biosensing platform to detect these viruses locally before outbreaks can occur and monitor their progression. To this end, we have developed an easy-to-use, cost-effective, multiplexed platform able to detect viral loads in wastewater with several orders of magnitude lower limit of detection than mass spectrometry. This is enabled by wafer scale production and aptamers pre-attached with linker molecules, producing forty-four chips at once. Each chip can simultaneously detect four target analytes using twenty transistors segregated into four sets of five for each analyte to allow for immediate statistical analysis. We show our platform’s ability to rapidly detect three virus proteins (SARS-CoV-2, RSV, and Influenza A) and a population normalization molecule (caffeine) in wastewater. Going forward, turning these devices into hand-held systems would enable waste-water epidemiology in low-resource settings and be instrumental for rapid, local outbreak prevention.

## 1. Introduction

According to the World Health Organization, lower respiratory infections are the fourth leading cause of death worldwide and second in low-income countries (World Health Organization 2014). The top three causes for these infections are SARS-CoV-2, Influenza, and Respiratory Syncytial Virus (RSV) (Madhi et al. 2020; Rouzé et al. 2021; Lafond et al. 2021). There is a growing emphasis on wastewater-based epidemiology (WBE) to track outbreaks. However, WBE is predominantly performed in high-income countries and densely populated areas (Ahmed et al. 2020; Hart and Halden 2020; Medema et al. 2020). Furthermore, if detection can occur on site, WBE would be instrumental to mitigating and tracking outbreaks from these viruses via early detection of viruses and other pathogens shed by asymptomatic carriers without requiring invasive and frequent individual tests (Champredon and Vanrolleghem 2023). For example, SARS-Cov-2 RNA can be detectable in wastewater 5 – 8 days before symptom onset and 2 – 4 days before positive clinical PCR tests (Peccia et al. 2020; Nemudryi et al. 2020).

Indeed, several college campuses exploited existing testing infrastructure to employ highly localized wastewater testing to prevent outbreaks during the Covid-19 pandemic. An instructive example is the University of California San Diego (UCSD), where sampling from 239 buildings across their campus allowed early hot spot detection and individual testing on a per-building basis (Karthikeyan et al. 2021, 2022). UCSD diagnosed nearly 85% of all SARS-CoV-2 infections on campus early and implemented preventative measures to mitigate the spread of the virus (Karthikeyan et al. 2021). This localized approach to WBE could also benefit low– and middle-income countries, where sewage is typically collected in individual or partially shared reservoirs (Street et al. 2020) that are not connected to community sewage systems (Adelodun et al. 2020). This is particularly relevant to RSV, a leading cause of respiratory-related deaths in those 0 – 5 years old (CDC 2023), where data from low– and middle-income countries is lacking or missing altogether due to inadequate systems and infrastructure needed to track disease transmission (Pawar 2023). Even in high-resource settings, the typical collection at a central waste-water facility limits sensitivity of pathogen detection in wastewater due to short half-lives of analytes of interest (Hart and Halden 2020) and natural dilution (Lowry, Wolfe, and Boehm 2023) of target biomarkers.

Several factors have hindered the widespread adoption of WBE and led to the general reliance on sample collection at centralized treatment facilities. Specifically, WBE testing is performed almost entirely utilizing advanced techniques in analytical chemistry and molecular biology, including liquid chromatography-mass spectrometry (LC-MS), high-pressure liquid chromatography-mass spectrometry (HPLC-MS), digital polymerase chain reaction, or real-time quantitative polymerase chain reaction (RT-qPCR) that requires dedicated lab space, personnel, equipment, and chemicals (Lorenzo and Picó 2019). Limited testing facilities and the need for sample collection and transport can also delay results and response times, limiting WBE for effective outbreak prevention (Leung 2021). Indeed, the WBE company BioBot in Cambridge, MA, says their average testing time is 11 – 15 days due to the need to test from multiple districts in weekly batches, creating a sample testing backlog (Biobot 2023b). Due to dilution of fecal waste in municipal wastewater, LC-MS and RT-qPCR rely on filtering and concentrating the collected sample (Adams 2020; Li, Wnkui; Zhang, Ji; Tse 2013), with HPLC-MS also subjecting it to several high-pressure steps to separate constituent elements (Else et al. 2010; Metabolite et al. 2023). Thus, a low-cost, easy-to-use, multiplexed device is urgently needed to enable point-of-need WBE.

Particularly challenging is the need of a sensing platform to withstand the harsh wastewater medium while accurately and reliably distinguishing between the various components. Wastewater can contain viruses shed in human waste and other particles ranging from naturally occurring biomass, bacteria strains, and drug metabolites to pharmaceuticals (Massano et al. 2023). Similarly challenging is the need to multiplex assays or testing strategies to monitor multiple targets to reduce cost, time, and effort while addressing seasonal and population variations via normalization. For point-of-need WBE sensing, population normalization is crucial due to increased variability in dilution factors, such as per capita water use, stormwater inputs, etc., and viral shedding rates (Sweetapple et al. 2023; Rainey et al. 2023; C. Li et al. 2022a). This variability exacerbates the already challenging task of calculating the number of people infected based solely on the virus concentration in the wastewater sample. For example, depending on the level of infection, a person suffering from SARS-CoV-2 can excrete anywhere from 600,000 (N. Zhang et al. 2020) to 30,000,000 virions/L (Wölfel et al. 2020) of fecal matter.

To enable WBE at the local level, especially in low-resource and rural communities, it is helpful to look towards efforts in personalized health care. Substantial efforts have been made regarding sensing respiratory infections using lateral flow immunoassay (LFIA), low-cost PCR, and electronic sensors. Electronic sensors are potentially the most promising as they can simultaneously offer multiplexed, low-cost, high sensitivity detection with minimal human effort. Here, there is growing interest in graphene field effect transistors (GFET), which have shown the capability to detect everything from lead ions (Velusamy et al. 2022; Dong et al. 2023) to bacteria and oral disease biomarkers (Ping et al. 2016b; Gao et al. 2016; Kumar, Gray, et al. 2020), though few have shown multiplexing capabilities (Lu et al. 2022; Kumar et al. 2022; Kumar, Gray, et al. 2020). Nonetheless, only two groups, including ours, have demonstrated GFET’s use for detection of analytes in wastewater. For instance, a GFET recently detected cadmium ions in wastewater with a limit of detection (LOD) of 0.125 pM (H. Wang et al. 2023). Still, virus protein detection in wastewater with GFETs, let alone by a scalable fabrication method, has not been shown.

This work focused on developing wafer-scale fabrication of GFET devices for rapid, easy-to-use, low-cost, multiplexed, and population-normalized detection of respiratory viruses in wastewater at low levels of detection (LOD). To do so, we implemented a new probe strategy where aptamers, single-stranded oligonucleotides, are pre-attached to the linker molecule, removing the need for harsh solvents. This enhanced the device’s reproducibility, lowering filtration levels and producing better LOD. In addition, we have optimized the fabrication process to make 44 chips simultaneously on a four-inch wafer. The devices are tested using freshly collected waste-water samples to detect SARS-CoV-2 spike protein, Influenza A hemagglutinin, RSV glycoprotein, and caffeine for comparison with lab-based WBE methods.

## 2. Materials and Methods

### 2.1 Graphene Platform Development

#### 2.1.1 Graphene as a transducer

Graphene is particularly useful yet challenging as a transducer due to its extreme sensitivity to surface charges (Castro Neto et al. 2009; Ping et al. 2016a; Hwang et al. 2016). Nonetheless, graphene is biocompatible and can be prepared at wafer scale. Due to its zero-band gap, it has a well-defined Dirac point (charge neutrality point) where its valence and conduction bands meet. This produces a peak in resistance when the chemical potential reaches the Dirac point (Fig. 1). When biomolecules attach to the surface of the graphene, it is generally assumed charge is transferred to graphene either directly or from conformal changes in the probe (J. Li et al. 2021; Seo et al. 2020a). This enables quantification of the target concentration via a shift in voltage at which the Dirac point appears.

**Fig. 1.**
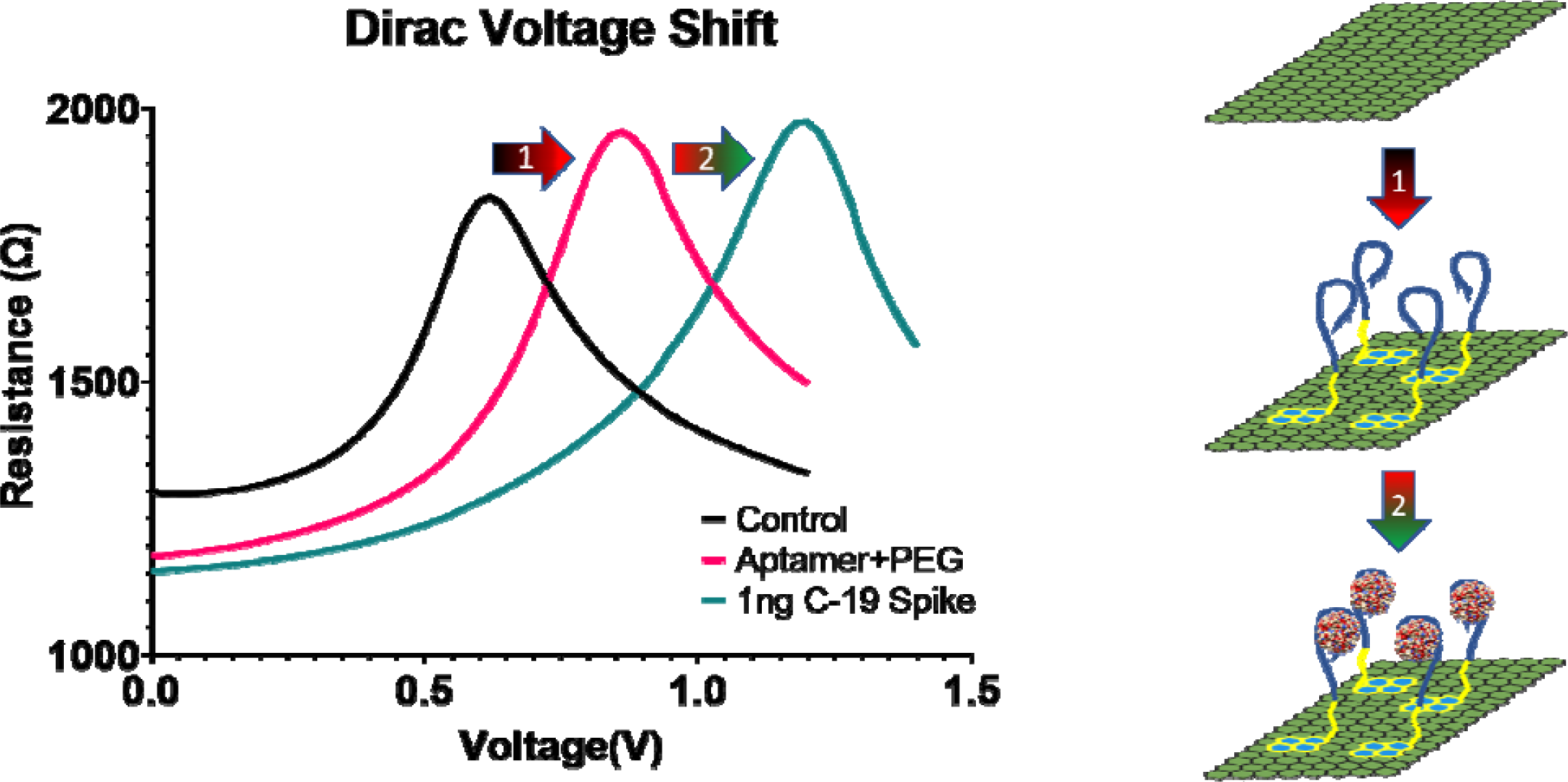
– Dirac voltage shifting with aptamer and target attachment: The plot on the left shows the positive shift in the Dirac point (peak of the curves) from the intrinsic position of the bare graphene (black) of approximately 0.6V. After a 2:1 mixture of the aptamer probe to PEG is added the Dirac point shifts positively to about 0.8V (pink). A large shift in the Dirac point to 1.2V is then seen (green) in the presence of 1ng/ml of the target protein for SARS-CoV-2. On the right is a schematic of the bare graphene, aptamer attachment, and target attachment.

Another advantage of graphene is the ease of functionalization with the biomolecules used as probes (Pinto, Gonçalves, and Magalhães 2013). These probes can be bonded to aromatic rings (e.g., Pyrene), which attach to the graphene through π-π stacking. This allows for tremendous biocompatibility between graphene and a host of biomolecules without unintentional disorder from chemical bonding. However, typically, graphene is functionalized via a two-step process, where the linker molecule is attached using dimethylformamide (DMF), and then the probe is later bound to the linker molecule (Seo et al. 2020b; Kwong Hong Tsang et al. 2019; https et al. 2022; Nekrasov et al. 2022). Unfortunately, the DMF tends to react with the device, causing instability, higher LOD, and lower reproducibility, and can attack polymers and passivation layers, degrading the device (Khan and Song 2021). As described later, we have avoided this issue and improved the LOD and reproducibility needed for point-of-need WBE using probes pre-attached to the linker molecule and incubated in PBS.

Likewise crucial for detection in complex wastewater matrices, graphene is insensitive to the sample medium’s pH levels (Fu et al. 2011). We demonstrated this in our recent work on opioid metabolite detection in wastewater (Kumar et al. 2022), in which we showed the simultaneous detection of Noroxycodone, Norfentanyl, and EDDP (2-ethylidene-1, 5-dimethyl-3, 3-diphenylpyrrolidine) with an LOD below that of HPLC-MS (Kumar et al. 2022). This work also exhibited our platform’s robustness and selectivity of the target molecules in wastewater. Unlike traditional field-effect transistor (FET) sensors that require large gate voltages (>60V) (Ping et al. 2016a), we have demonstrated our ionic liquid-gated GFETs are compatible with simple electronics requiring less than 2V (Kumar, Gray, et al. 2020; Kumar, Wang, et al. 2020).

#### 2.1.2 GFET Fabrication, Characterization, and Functionalization

To bring our GFET-based Graphene Electronic Multiplexed Sensor (GEMS) towards point-of-need WBE, we modified our fabrication method and device design to enable production on a four-inch silicon wafer (Fig. 2a – Top) before dicing into individual chips. This has drastically reduced our costs per chip primarily due to a drastic decrease in fabrication time. Prior to wafer-scale fabrication, we were able to produce 6 – 8 chips in four days. We can now produce 44 GEMS in the same amount of time. Each GEMS has 20 GFETs arranged in groups of five for rapid statistical analysis of variability between GFET devices. To enable multiplexed detection, the groups are segregated with PDMS wells with individual coplanar side gates (Fig. 2b). This enables individual functionalization of each well with a different probe without cross-functionalization.

**Fig. 2.**
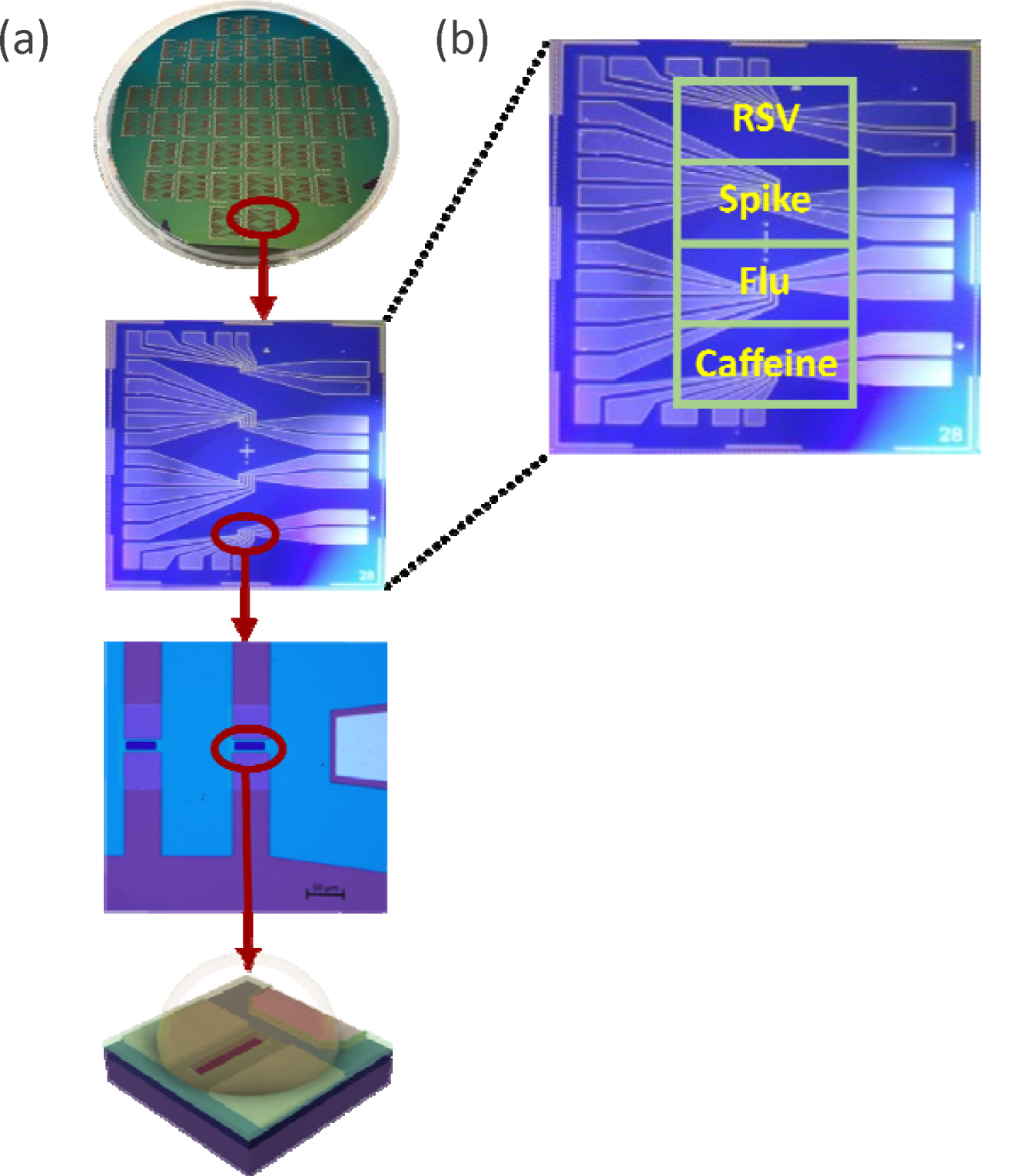
– (a) Top: Wafer as fully fabricated. Second from top: Overview of the 1.2cm x 1.2cm GFET sensing platform. Third from top: 20x microscope image of a single sensing well. Two graphene devices and the coplanar gate electrode are shown. Bottom: Diagram of a single, functionalized graphene device during the sensing process. (b) Schematic of individual chip with PDMS well and labeled with functionalization for specific analytes.

We first pattern bottom contacts on a four-inch Si/SiO_2_ wafer using bi-layer photoresist (LOR1A/S1805) and photolithography followed by e-beam deposition of 5 nm of titanium wetting layer and 20 nm of platinum. Platinum is chosen to minimize contact resistance to graphene because it is robust and has low surface potential (Fujii, Kasuya, and Kurihara 2017). After metal liftoff, the contacts were annealed under vacuum for 10 hours at 400°C to remove any remaining photoresist and increase the electrodes’ smoothness, allowing for better graphene attachment. CVD graphene was transferred on top of the entire wafer by General Graphene Corp. in Knoxville, TN. The wafer was then annealed under vacuum in the e-beam chamber for nine hours at 300°C to remove any remaining residues and water from the transfer process. Before removing from the e-beam chamber, 3 nm of aluminum oxide (AlOx) was deposited to protect the graphene from further chemicals and atmosphere during later fabrication steps. Once removed, the wafer was baked on a hotplate in our glovebox at 175°C for five minutes to ensure aluminum oxide adhesion. The same bi-layer resist process and photolithography system were then used to pattern the graphene for etching via oxygen plasma. The MF-321 developer (from Kayaku) used to develop the pattern after lithography has the added benefit of also removing the 3 nm of AlOx from atop the graphene we wish to etch. This was followed by argon plasma to remove any oxide layer formed on the platinum by the oxygen plasma on the coplanar side gate. Failing to remove this layer has led to higher initial Dirac points and, in turn, lower sensitivity in our devices.

Next, the devices were cleaned with Remover PG and rinsed with IPA and DI water. The chips were then baked under a vacuum at 200°C for one hour to remove any water and clean any residue from the wafer. After this, a 50 nm passivation layer of aluminum oxide was deposited to encapsulate the devices while the wafer was still hot. Oxygen was flowed to achieve a pressure of ∼10^5^ Torr during AlOx deposition to replenish oxygen stripped from the AlOx crystals during e-beam deposition. A final single layer (S1805) photolithography step was then performed to expose the graphene sensing windows (10um x 40um) and the contact pads for wire bonding. Exposed AlOx was then etched with 65:35 diluted TRANSETCH-N (from Transene) for 14 minutes at 80°C, then rinsed with DI water. The remaining photoresist was then removed with Remover PG and rinsed with IPA and DI water. The wafer was then diced using a Pelco Wafer Dicing system, eliminating the need for a wafer dicing saw and its associated chemicals. The chips were then mounted to chip carriers and wire-bonded. Following this, PDMS wells made in-house with custom 3D-printed molds were placed on the chips to hold the functionalization liquids and target mixtures during incubation as per our sensing protocols.

### 2.2 Pre-Linked Aptamers

We employ aptamer probes due to their high affinity, stability, and small size (Cai et al. 2018; Urmann et al. 2017). Aptamer-based protein biosensing depends on aptamer-target binding (Y. Wu et al. 2014), which several factors can complicate. Structurally complex protein targets have more binding sites and interaction types than small molecules (S. Jones et al. 2001; Kohlberger and Gadermaier 2022).

This increase in complexity can result in aptamers with decreased target specificity if the experimental design of SELEX is flawed (Qian et al. 2022). Generation of aptamers for proteins via SELEX is more manageable for small molecules (Y. Wu et al. 2014), but the conformation of the protein (purified or native) can alter or hinder aptamer binding (Zhou and Rossi 2017; Z. Zhang et al. 2021). Careful consideration is necessary to ensure binding conditions mirror real-world binding conditions. With this in mind, we chose the Universal Aptamer (UA) (Bhardwaj et al. 2019; Shiratori et al. 2014; C.-H. Wang, Chang, and Lee 2016) for Influenza A hemagglutinin, H8 (Percze et al. 2017) for RSV, and 1C (Y. Zhang, Juhas, and Kwok 2022) for SARS-CoV-2 spike proteins based on their binding affinities to their targets. See the Supplemental for further information regarding the aptamers.

Generally, to attach the aptamer to graphene, the device is first incubated with 10 mM 1-pyrenebutyric acid N-hydroxysuccinimide ester (PBASE) linker molecule dissolved in DMF for one hour. After performing a Dirac point measurement to see the shift due to DMF and PBASE, a 2:1 mixture of aptamer to polyethylene glycol (PEG) is incubated for one hour. Adding PEG to the probe mixture has been widely employed (Szunerits et al. 2023; Rodrigues et al. 2022) to prevent unwanted attachment of molecules to any unlinked PBASE molecules and provide space between aptamers, limiting their interactions. The PEG also stabilizes the devices by minimizing drift and standard deviation between different devices.

To further reduce cost, analysis, and fabrication time and boost reproducibility, we altered this typical process by pre-attaching the aptamers and PEG to the PBASE molecules (See supplementary information for more details regarding pre-linking.). Indeed, DMF is known to dope graphene (G. Wu et al. 2017) and thus would be expected to result in a higher limit of detection. With this in mind, we performed identical experiments with GEMS using the standard DMF attachment procedure and our pre-linked PEG (PL-PEG) and probes (PL-aptamers). As seen in Fig. 3 and Table 1, more significant Dirac point shifts occurring at much lower LODs are seen in devices with pre-linked (PL) probes. Lastly, to ensure graphene cleanliness and device reproducibility, much of the fabrication was carried out in a pure argon environment inside our cleanroom-in-a-glovebox (Gray et al. 2020).

**Fig. 3.**
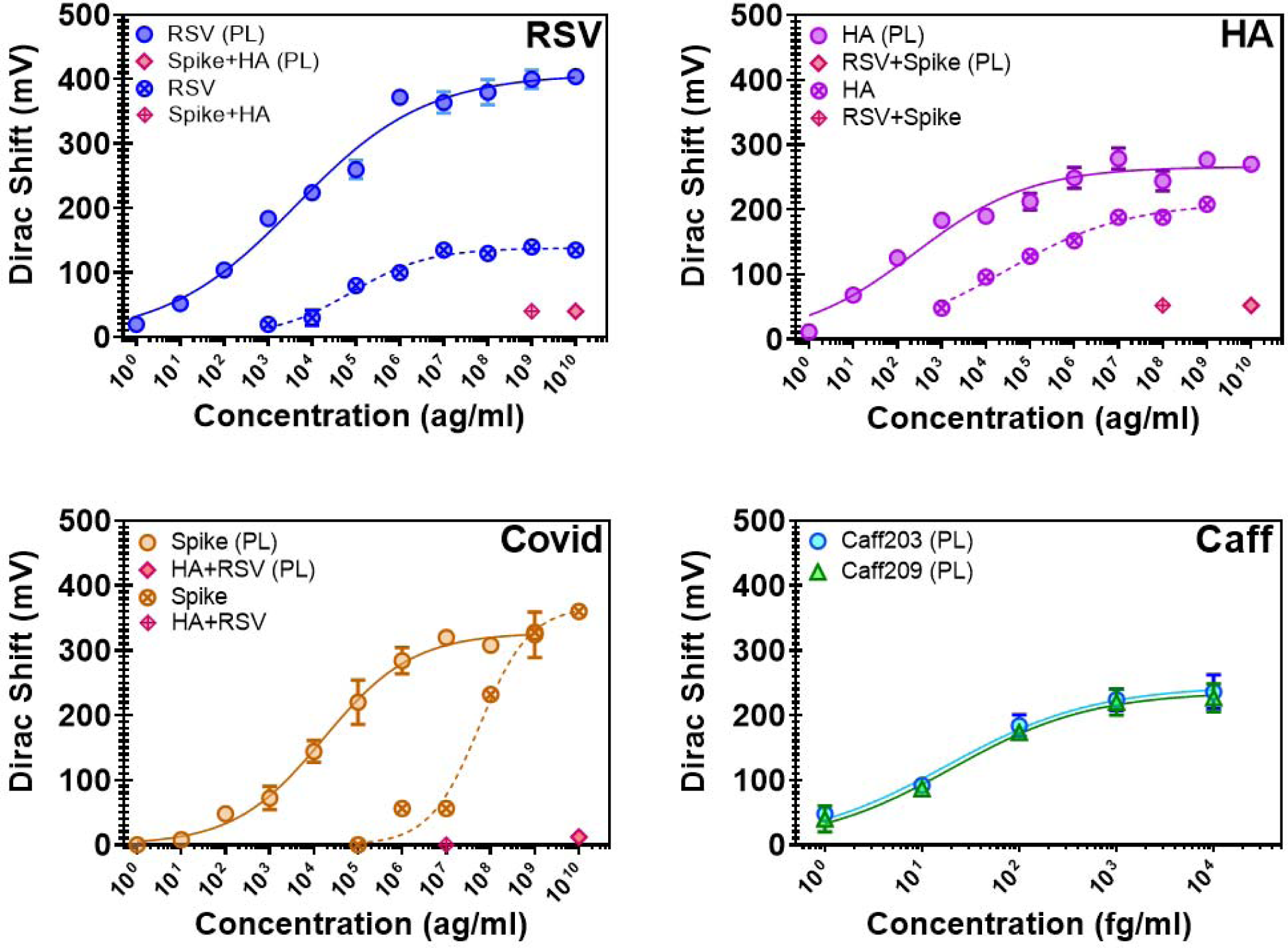
–Concentration dependance measurements of viral proteins in PBS. Error bars calculated from the five GFETs per sensing well. Data points with crosses and dashed curves indicate non-pre-linked aptamers were used. Top Left – Influenza A Hemagglutinin detection. High concentrations of RSV and COVID Spike proteins used as a negative control. (PL) denotes pre-linked aptamer experiment. Top Right and Bottom Left – Same as in HA plot but with SARS-CoV-2 Spike and RSV proteins, respectively. Non-target proteins used as negative control in each case. Bottom Right – Concentration dependence measurements of two caffeine aptamers. Caffeine measurements were only conducted with pre-linked aptamers.

**Table 1.** Comparison of LODs between pre-linked and unlinked aptamers for each target analyte in PBS. All pre-linked virus experiments were conducted on a single GFET chip and unlinked on another. Caffeine experiments were only performed with pre-linked aptamers and done on a single GFET chip.

| Target | Unlinked Aptamer | Pre-Linked Aptamer |
| --- | --- | --- |
| SARS-CoV-2 Spike Protein | 91 pg/ml | 55 ag/ml |
| Hemagglutinin (Flu A) | 79 fg/ml | 408 ag/ml |
| Respiratory Syncytial Virus Protein | 43 fg/ml | 453 ag/ml |
| Caffeine | N/A | Caff203: 35 fg/ml<br>Caff209: 26 fg/ml |

### 2.3 Optimization in PBS

#### 2.3.1 Viral aptamers

Selectivity and concentration analysis was first conducted in 1x PBS to determine the aptamer viability without the background signal from wastewater. Specifically, the numerous constituent components in wastewater (Henze, Mogens; van Loosdrecht, Mark; Ekama, George; Brdjanovic 2008; M. H. Huang, Li, and Gu 2010; Novo et al. 2013), many of which are charged ions, can produce false positives. For each target, we first determined the initial Dirac point of the graphene in 0.01x PBS (see Supplemental). Diluted PBS minimizes the Debye screening effect (Stern 2007). Typically, we observe a Dirac point around 0.6 V (±0.1V) due to the work function of the platinum side gate electrode (Fujii, Kasuya, and Kurihara 2017). This baseline Dirac point ensures the graphene quality without unwanted doping. This is further confirmed by the nearly symmetric slopes to the left (hole regime) and to the right (electron regime) of the Dirac point, which results from the charge carrier mobilities (Gosling et al. 2021). Passivation issues are typically indicated by double peaks in the curves. Good passivation is also confirmed by ensuring the Dirac point does not drift with repeated gate voltage sweeps. The highest quality devices have an initial Dirac point in the range of 0.58 – 0.7 V with an average starting resistance around 2000 Ω and a stable Dirac point after three measurements. Data on initial Dirac point and starting resistances were collected for 545 different GFETs fabricated over two years in our lab, showing that most of our devices fall within these parameters (see Supplemental).

After initial testing, we incubated the graphene devices for one hour with a 2:1 mixture of 10uM PL-aptamer to 10uM PL-PEG, which was optimized in our previous work with opioids in wastewater and oral disease biomarkers in saliva (Kumar, Wang, et al. 2020; Kumar, Gray, et al. 2020; Kumar et al. 2022). Dirac point measurements are again conducted in 0.01x PBS to confirm attachment to the graphene surface. Upon attachment, the charged phosphate backbone of the aptamer induces positive charge carriers into the graphene, producing a positive 150-200mV shift in the Dirac point (see Supplemental). Atomic force microscopy and Raman measurements have also been performed to confirm the attachment (see Supplemental).

We first assessed all aptamer selectivity against a negative control. For example, the Influenza A hemagglutinin (HA) with a concentration of 10 – 100 ng/ml that is far beyond that found in wastewater (tens of pg/ml), is incubated on the devices for one hour in the well containing the SARS-COV-2 SPIKE PROTEIN aptamer (1C). No shift in the Dirac point was seen, showing the HA protein does not bind to the 1C aptamer (Fig. 3 – Covid). Similar negative control analyses were conducted in the wells functionalized with the Influenza and RSV aptamers. As shown in Fig. 3, these aptamers had a slightly higher non-specific interaction with the negative control proteins. Nonetheless, the Dirac point shifts in wells with the Influenza and RSV aptamers resulting from negative controls were relatively small (approximately 50 mV), setting the baseline for future measurements.

Next, we focused on assessing each aptamer’s limit of detection and affinity. We followed a standard protocol of incubating the devices with a specific concentration of the target proteins. After incubation, the device is rinsed with 1x PBS and DI water before performing the Dirac point measurement in 0.01x PBS. For each concentration, the reported shift is the difference in the Dirac point value obtained from that of the negative control. After measuring the Dirac shift, we incubated with increasing target protein concentrations. To ensure the absence of systematic errors, we have also performed measurements with random concentrations to ensure they match the signal detected by a systematic increase in concentration.

Beginning with low concentration, each incubation is conducted for one hour. We found that concentrations below 1 fg/ml for the SARS-COV-2 spike protein did not change the Dirac point. However, the RSV and HA proteins produced shifts at much lower concentrations (approximately 10 ag/ml). This shift discrepancy may be due to the newness of the SARS-COV-2 spike aptamer and future improvements can improve its binding affinity. The average shift from all devices in the well and their standard deviations are plotted in the same graph as the negative control’s shift. The concentrations are increased by one order of magnitude in each subsequent incubation, and the same rinsing and sensing protocol is conducted for each. The concentrations are increased until a saturation point is reached, determined by no further shift with two consecutive high concentrations.

Upon collecting the concentration dependence, we found the binding characteristics of the aptamer by fitting the Dirac voltage shift versus target analyte concentration to Hill’s equation (Goutelle et al. 2008):

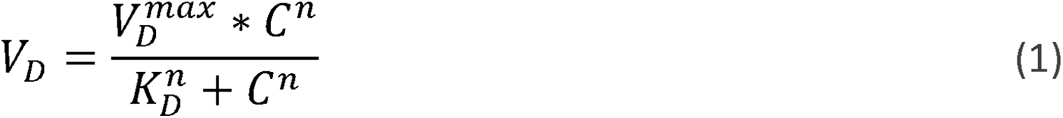

Here, *V_D_* is the Dirac voltage shift measured in mV, 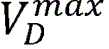 is the maximum Dirac voltage shift at the saturation point, *C* is the concentration of the target analyte, *n* is the Hill Coefficient determined to be the maximum slope on a log plot of the response curve, and *K_D_* is the dissociation constant. The parameters were found using a least squares fit model in Matlab after providing estimates of the Hill Coefficient, maximum Dirac voltage, and dissociation constant. Due to the five devices in each well of the GFET, we can perform statistical analysis immediately. This allows us to calculate the LOD for each analyte by using the residuals of the standard deviation against the Hill fit using 3σ analysis (Belter, Sajnóg, and Barałkiewicz 2014):

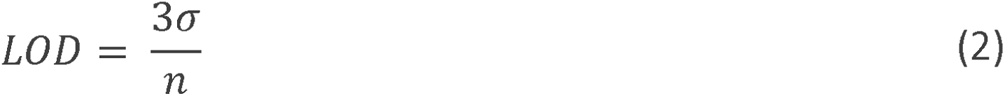

Here, σ is the standard deviation from the fit and *n* is again the Hill slope. This was used to find the LODs in Table 1.

#### 2.3.2 Caffeine aptamer

WBE programs use several different biomarkers to determine the total contributing population. These include caffeine, paraxanthine (caffeine’s metabolite), creatine, 5-hydroxyindoleacetic acid (5-HIAA, serotonin metabolite), and pepper mild mottle virus (PMMoV) given their ubiquitousness in human diets and survivability in wastewater (Hsu et al. 2022). Paraxanthine and PMMoV concentration, in particular, are excellent means for population normalization (C. Li et al. 2022b). Unfortunately, to the best of our knowledge, no aptamer has yet been developed for PMMoV or paraxanthine. Therefore, to test our platform’s capabilities as a means for population normalization in wastewater, two previously reported caffeine aptamers were selected based on their reported results that show micromolar sensitivity in human serum (P. J. J. Huang and Liu 2022), two of which (Caff203 and Caff209) were chosen for our tests in wastewater. Both the Caff203 and Caff209 aptamers were pre-attached to the PBASE linker molecules, and the same functionalization and sensing protocols were followed as the virus proteins. Both were evaluated first in PBS to determine their viability before exposure to wastewater. Caff203 was found to have an LOD of 35 fg/ml in PBS, while Caff209 showed 26 fg/ml in PBS (Fig. 3). Due to its lower LOD, Caff209 was selected for future experiments.

## 3. Results

### 3.1 Wastewater biosensing

#### 3.1.1 Wastewater dilution optimization

Next, we turned to testing GEMS with wastewater. In our earlier work on opioid metabolites, we found diluting the wastewater with 1x PBS to a 20:1 mixture necessary to minimize unwanted Dirac point shifts and false positives from the myriad components and non-neutral pH (6-9). Given the improved device performance with pre-attachment, we re-optimized this dilution to attempt a lower LOD. We began by incubating the 1C PL-aptamer and PL-PEG, as previously discussed. The wastewater was then passed through a 0.3-micron filter to remove large particulates. Next, various dilutions (2:1, 5:1, 10:1, and 20:1) were incubated directly on the devices for one hour, and the resulting Dirac point shift is shown in Fig. 4. We found the 10x dilution caused an approximate 60 mV Dirac point shift, the same as the 20x dilution. Since PBS does not induce a shift, the background signal from wastewater will increase the LOD by setting a floor below which we cannot uniquely detect the target, as indicated by the horizontal dashed lines in Fig. 5. Next, four samples were diluted with the 2:1, 5:1, 10:1, and 20:1

**Fig. 4.**
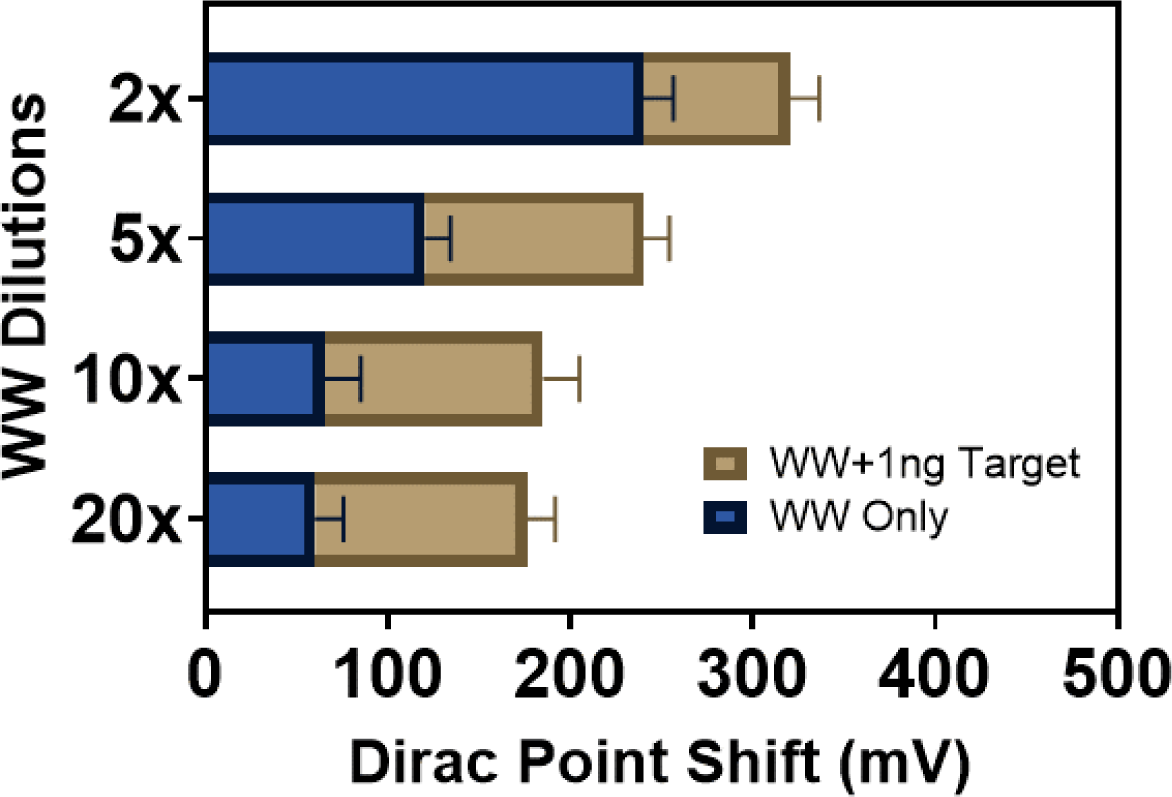
– Histogram of the average Dirac point shift at various wastewater dilutions. The blue areas show the average Dirac point shift for five GFET devices after incubation of diluted wastewater for one hour. The tan areas show the further Dirac point shift after incubating the GFETs for one hour with 1ng/ml of target protein in their respective wastewater dilutions.

**Fig. 5.**
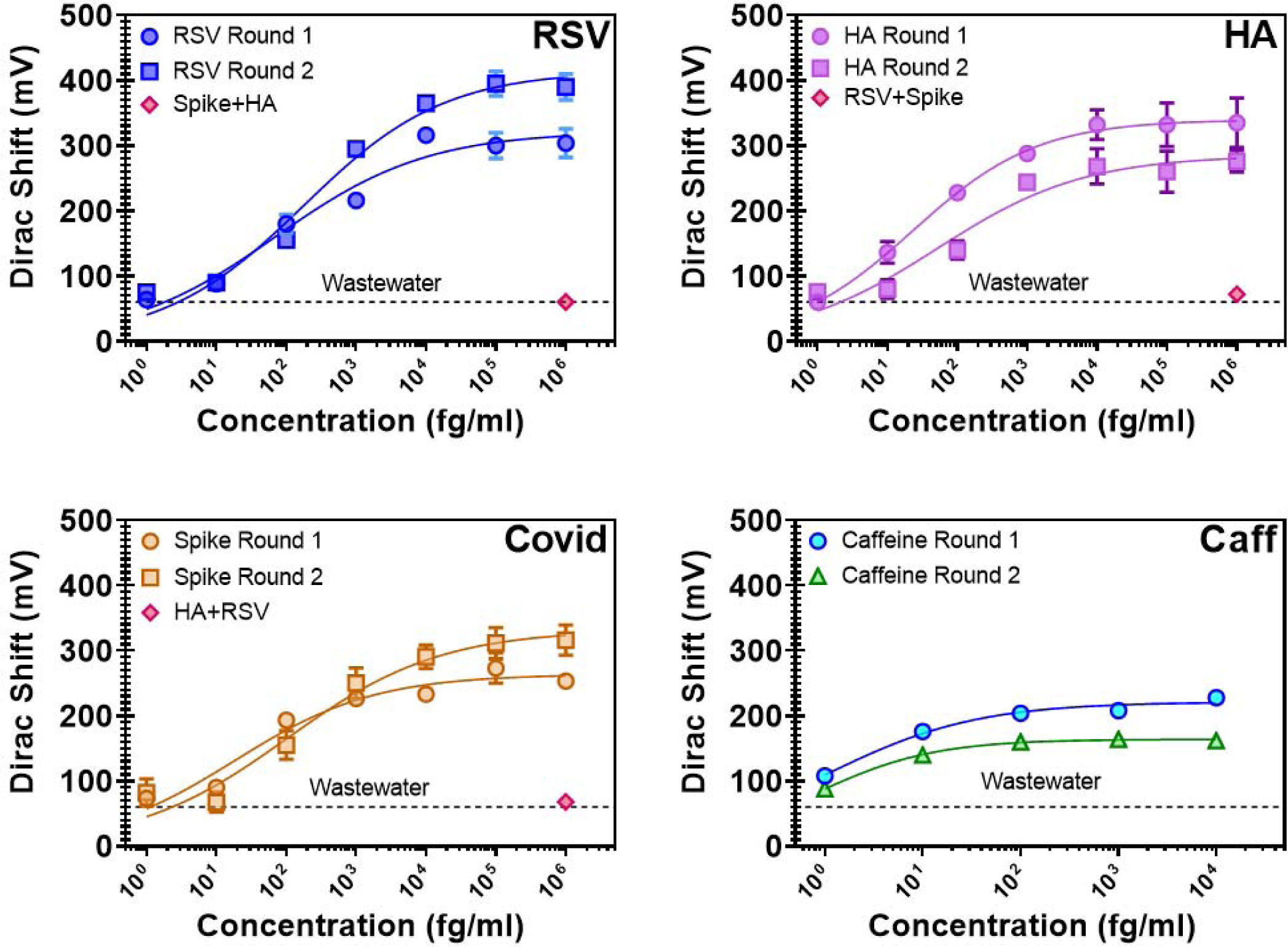
– Concentration dependence measurements of viral proteins and caffeine in wastewater. Error bars calculated from the five GFETs per sensing well. From left to right, HA, RSV, and COVID spike proteins and Caffeine. Horizontal dashed line shows intrinsic background shift from the wastewater itself.

PBS to wastewater samples to create 1 ng/ml solutions of SARS-COV-2 spike protein and incubated on the devices for one hour. This was done to determine if wastewater dilution affected the ability of the aptamers to find the target proteins. The 1 ng/ml concentration was used since this is the point at which the SARS-COV-2 SPIKE PROTEIN aptamer saturated when tested in 1x PBS. Interestingly, there was a statistically insignificant difference in the shift between the 10:1 and 20:1 wells. Both measured a shift of around 130 mV after incubating with the spike protein mixture. Thus, we focused 10:1 PBS to wastewater dilution to achieve the smallest possible LOD in wastewater.

#### 3.1.2 Detection of Analytes in Wastewater

Having optimized the wastewater dilution, we performed a similar series of concentration-dependent measurements with the same protocol done first in PBS. The experiments were conducted in two rounds for each analyte. Experiments were first performed on a single GFET chip. Four wells were functionalized with a different pre-attached aptamer: 1C for SARS-CoV-2, UA for hemagglutinin, H8 for RSV, and Caff209 for caffeine. A fresh wastewater sample was obtained (collected one day prior and stored a 4°C overnight), filtered, and diluted in a 10:1 ratio with PBS and spiked with virus proteins and caffeine to make concentrations ranging from 1 fg/ml to 1 ng/ml with an increase of one order of magnitude between each concentration. The second round of experiments was conducted one month later using a newly fabricated GFET chip, fresh pre-attached aptamers, and a new wastewater sample. In both instances, the negative controls were tested first at 1 ng/ml to check selectivity, followed by increasing the concentrations of the target analyte. In both rounds, the negative controls showed little to no shift beyond the background 60mV shift from the wastewater (dashed lines in Fig. 5).

The resulting concentration curves are shown in Fig. 5, and LODs for each round are shown in Table 2. As expected, LOD values increased over the PBS results due to the intrinsic 60mV signal from the wastewater. Nonetheless, the larger LODs are all well within the range for the concentrations of each analyte in wastewater. SARS-CoV-2 has been shown to contain 24±9 spike proteins per virion (Ke et al. 2020), theoretically suggesting the LOD for our GEMS platform to be on the order of 27 – 59 virions/ml (27,000 – 59,000 virions/L) in wastewater assuming fully lysed virions. Influenza A has been found to contain 300 – 400 HA proteins per virion (Einav, Gentles, and Bloom 2020), giving a theoretical LOD of fully lysed virions in the 1.5 – 7 virions/ml (1,500 – 7000 virions/L). To the best of our knowledge, the average number of proteins for RSV has not yet been determined. Assuming a similar number between the spike and the Influenza A proteins, the theoretical, fully lysed RSV virions could be 15 – 397 virions/ml (15,000 – 397,000 virions/L). Like the experiments conducted in PBS, the RSV and SARS-COV-2 spike aptamers show little to no shift with the high concentration of negative control. In contrast, the HA aptamer showed a small but significant shift of around 60 mV with negative control. This could be partly due to UA’s longer length compared to the others, allowing it to bind to more constituent elements in the wastewater. It could also be due to HA proteins already present in the wastewater sample, which was collected during the 2022 – 2023 Flu season.

**Table 2.** Limits of Detection (LOD) for each target analyte from two separate experimental rounds. Each round was conducted on a single GFET chip. Based on their average molecular weights, LODs were converted from fg/ml to proteins/ml.

| Target | Round 1 LOD | Round 2 LOD | Reported LC-MS from Literature |
| --- | --- | --- | --- |
| SARS-CoV-2 Spike Protein | 136 fg/ml<br>(890 proteins/ml) | 187 fg/ml<br>(1224 proteins/ml) | $10^5 - 10^6$ copies/ml (Griffin and Downard 2021; Dollman, Griffin, and Downard 2020; Nikolaev et al. 2020; Picó and Barceló 2021)<br>(~3000 fg/ml) |
| Hemagglutinin (Flu A) | 39.6 fg/ml<br>(612 proteins/ml) | 181 fg/ml<br>(2799 proteins/ml) | $7 \times 10^6$ copies/ml (Bojórquez-Velázquez et al. 2022)<br>(~30,000 fg/ml) |
| Respiratory Syncytial Virus glycoprotein | 176 fg/ml<br>(5927 proteins/ml) | 175.4 fg/ml<br>(5959 proteins/ml) | $3.6 \times 10^7$ copies/ml (Bojórquez-Velázquez et al. 2022)<br>(~40,000 fg/ml) |
| Caffeine | 10 fg/ml<br>($3.1 \times 10^7$ molecules/ml) | 2 fg/ml<br>($6.2 \times 10^6$ molecules/ml) | 5000 fg/ml (Huerta-Fontela, Galceran, and Ventura 2007) |

Due to its lower LOD found in PBS (Table 1), Caff209 was selected for analysis in wastewater. Interestingly, the LOD in wastewater was lower than in PBS, which was not seen with the virus proteins. This could be due in part to the salt content in wastewater facilitating binding (Lores and Pennock 1998) of the much smaller caffeine molecules, which are 0.194 kDa as compared to the larger proteins having sizes of 139.7 kDa, 59 kDa, and 37 kDa for spike, HA, and RSV respectively, lowering the variability between the devices.

To put these LODs in context, we compare them with other reported LODs. BioBot reports a limit of detection (LOD) for SARS-CoV-2 of 9000 copies/L using RT-qPCR (Biobot 2023a) (approximately 10 whole virions/L), which is lower than concentrations typically found in wastewater. The reliance on lab testing results from these low virus loads in wastewater requires amplification and/or viral concentration steps to detect. These concentrations can range from, in the case of SARS-CoV-2, 150,000 – 141.5 million viral genome copies (150 – 141,500 whole virions (Sender et al. 2020)) per liter of wastewater (Hart and Halden 2020). Influenza A concentrations are reported to be around 260,000 copies per liter (Heijnen and Medema 2011) and RSV 1,071 – 70,700 copies per liter (Ahmed et al. 2023). Others have reported LODs from RT-qPCR as low as 2.9 – 4.6 copies per reaction after concentrating the sample from 50ml to 20ul (Ahmed et al. 2022). Several studies have found levels of shed virus can vary substantially depending on patient infection level and virus variants, ranging from 10^2^ – 10^7^ copies/ml (D. L. Jones et al. 2020; Pan et al. 2020; Zang et al. 2020; Han et al. 2020). These levels will significantly decrease upon reaching a wastewater treatment facility due to dilution and virus decay, highlighting the need for more localized collection and analysis. While our GEMS platform cannot achieve the low LODs seen with RT-qPCR, its LODs are 1 – 2 orders of magnitude lower than what has been reported with LC-MS (Table 2).

In the case of LC-MS, LODs between 10^5^ – 10^6^ copies per nasopharynx sample (Dollman, Griffin, and Downard 2020; Nikolaev et al. 2020) have been reported. While LC-MS has been used to detect SARS-CoV-2 in wastewater (Peng et al. 2022; Lara-Jacobo et al. 2022), no detection limit has yet to be reported. So-called “rapid tests,” on the other hand, while having short analysis time, typically rely on LFIA, which to the best of our knowledge have not shown the ability to rapidly sense the low level of virus in unprocessed wastewater. An LFIA sensed human adenovirus in processed wastewater by first concentrating the wastewater sample through PEG precipitation overnight and then performing a recombinase polymerase amplification step and achieving an LOD of 50 copies/reaction starting from an initial sample size of 1L (Rames and Macdonald 2019). While this is a low LOD, the tradeoff is in the time and complexity of the analysis.

### 3.2 Blind Testing

To ensure device integrity, a blind test in wastewater was performed. A single chip was functionalized with each virus aptamer in a different well. Four concentrations of each target protein were made by one author (O.R.P.) and were coded with a four-digit number (1738, 1993, 2930) with no indication of the contents. These were tested by another author (M.G.) using the same sensing protocol outlined above to determine which coded sample contained each protein. As shown in Fig. 6, for concentrations above 100 pg/mL (consistent with our earlier LOD) the target can easily be identified by only producing a Dirac shift in one well. Based on this, M.G. identified each target, which was confirmed correct by O.R.P.’s written records.

**Fig. 6.**
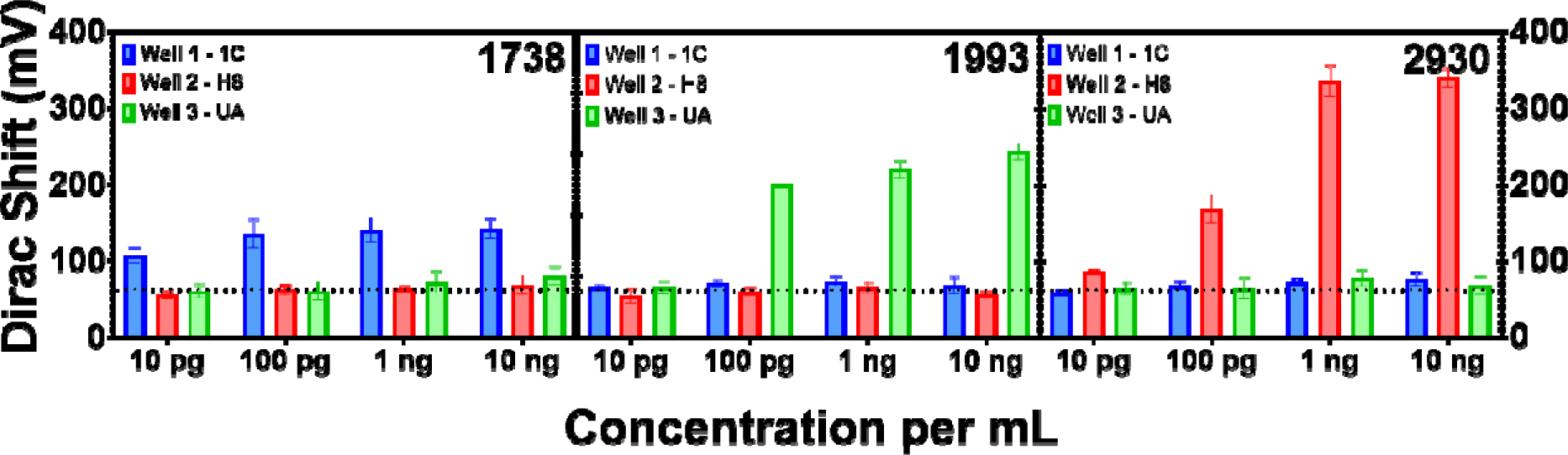
– Wastewater Blind Tests: Each plot represents the differing concentrations for a coded sample. The assorted colors indicate the aptamer used in each well; blue for the 1C (Covid), red for H8 (RSV), and green for UA (Flu). The horizontal dotted line is the intrinsic shift from the wastewater. M.G. found that 1738 was COVID spike proteins (left), 1993 was HA (middle), and 2930 was RSV. Each was confirmed by O.R.P.’s written records.

## 4. Conclusions and Future Work

To summarize, we showed the viability of our GEMS platform for selective, specific, simultaneous, and highly sensitive detection of four different analytes in wastewater, including caffeine for population normalization and three different viral proteins. We achieved limits of detection (see Table 1) one to two orders of magnitude better than HPLC-MS (Fig. 5) (Dollman, Griffin, and Downard 2020; Nikolaev et al. 2020; Peng et al. 2022; Lara-Jacobo et al. 2022; Kasprzyk-Hordern et al. 2023; Mestankova et al. 2012) and below the levels needed for effective early interventions (Peng et al. 2022). Results are obtained using a 1 cm^2^ chip in just over one hour with minimal human intervention and without bulky, expensive lab equipment or costly reagents. Simple wastewater preparation can be easily performed with minimal training, while low voltage and resistance ranges can be operated with simple and cheap electronics. The cost is minimized by wafer-scale fabrication and pre-linked aptamers, further enhancing reproducibility and LOD. Combined with our previous results, the scalable GEMS platform enables rapid, easy, and cheap wastewater sensing of a wide range of analytes (opioid metabolites, viruses, etc.). This shows our platform to be a practical choice for wastewater-based epidemiology for viral testing and can lead to finding hotspots for future virus outbreaks. Our platform’s low cost and power requirements could allow WBE to be performed on a building-by-building level in low-resource or rural settings, ushering in a new era of wastewater testing. Enabling this will require future efforts for on-chip electronics and microfluidics for sample preparation and a more comprehensive array of analytes to be tested on the same chip.

## Author Information

### Corresponding Author

Kenneth S. Burch − Department of Physics, Boston College, Chestnut Hill, Massachusetts 02467, United States; orcid.org/0000-0002-7541-0245;

### Authors

Michael Geiwitz – Department of Physics, Boston College, Chestnut Hill, MA 02467, United States; orcid.org/0009-0000-7197-9381

Owen Rivers Page – Department of Biology, Boston College, Chestnut Hill, Massachusetts 02467, United States; orcid.org/0000-0003-4072-8509

Tio Marello – Department of Physics, Boston College, Chestnut Hill, MA 02467, United States

Narendra Kumar – GRIP Molecular Technologies, Inc., 1000 Westgate Drive, Saint Paul, MN 55114; orcid.org/0000-0002-5319-1547

Stephen Hummel – Department of Chemistry and Life Science, United States Military Academy, West Point, NY 10996, USA

Vsevolod Belosevich – Department of Physics, Boston College, Chestnut Hill, MA 02467, United States

Qiong Ma – Department of Physics, Boston College, Chestnut Hill, MA 02467, United States

Tim van Opijnen – Department of Biology, Boston College, Chestnut Hill, Massachusetts 02467, United States

Bruce Batten – GRIP Molecular Technologies, Inc., 1000 Westgate Drive, Saint Paul, MN 55114

Michelle M. Meyer – Department of Biology, Boston College, Chestnut Hill, Massachusetts 02467, United States; orcid.org/0000-0001-7014-9271

### CRediT authorship contribution statement

K.S.B., M.G., and N.K. conceived the project and designed the experiments. M.G. fabricated, functionalized, and tested all GFET platforms. M.G., N.K., S.H, T.v.O., and K.S.B. selected all aptamers and analyzed the data. O.R.P. pre-linked all aptamers with PBASE linker molecules. T.M. aided in the design and placement of PDMS wells. S.H. validated 1C aptamer with flow cytometry. V.B. performed AFM and wire bonding machine maintenance. Q.M. supervised AFM and wire bonding maintenance. T.v.O. supervised flow cytometry. M.M. supervised aptamer pre-linking and blind test sample preparation. B.B. and K.S.B. aided in data analysis. M.G., O.R.P., and K.S.B. wrote the manuscript.

## Data Availability

All data is available on request.

## Acknowledgements

The work of M.G., T.M., and K.S.B. are grateful for the support of the National Science Foundation (NSF) EPMD program via grant 2211334. M.M.M., K.S.B. and O.P are grateful for support from the Boston College Schiller Institute for Integrated Science and Society Exploratory Collaborative Scholarship grant. M.G. would like to thank the Boston College Cleanroom & Nanofabrication Facility’s staff, Stephen Shepard and Dr. Chris Gunderson. M.G. and K.S.B. also thank Dr. Catherine Hoar for her insights on WBE. We would also like to thank Mr. Bryan Horsely, Mr. George Heufelder, and Dr. Sara Wigginton from MASSTC for providing wastewater samples and analysis of their source wastewater contents. Q.M. and V.B. acknowledge the American Chemical Society Petroleum Research Fund (PRF) 66299-DNI10.

