## Supplemental for "Graphene Multiplexed Sensor for Point-of-Need Viral Wastewater-Based Epidemiology"

**S1.** Concentration curves from non-prelinked aptamers in wastewater

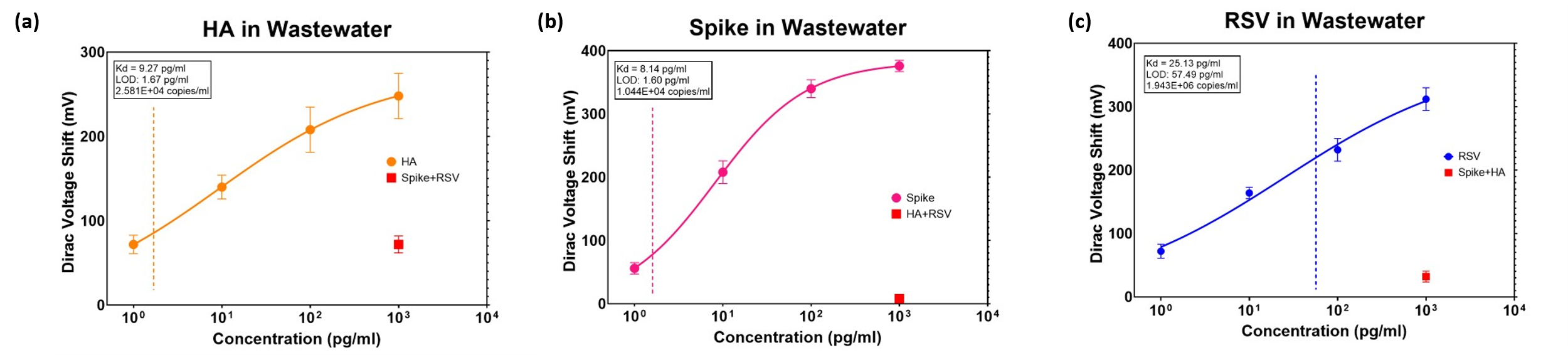

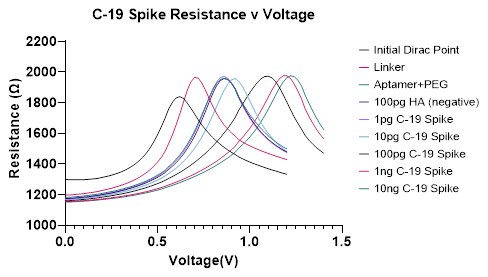
**S2.** Example Resistance vs. Voltage curves showing Dirac point shifts during experimentation.

**S3.** Reproducibility analysis from over two years of device fabrication. Total number of devices is N = 545

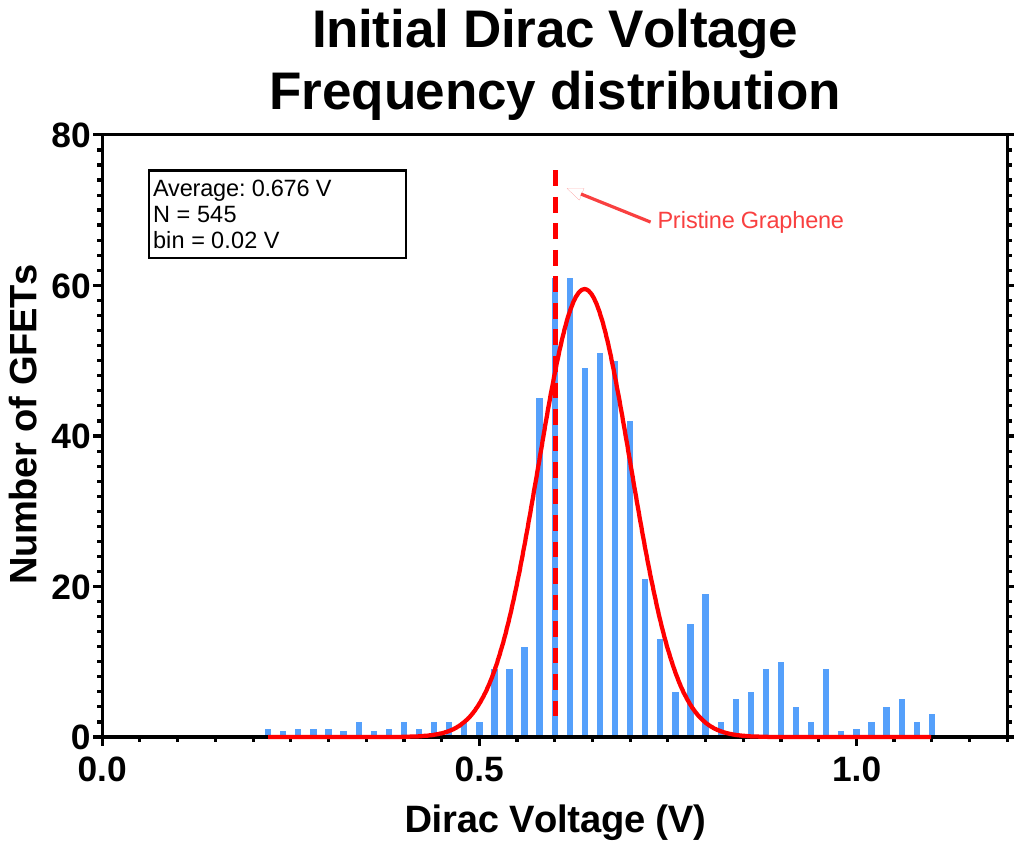

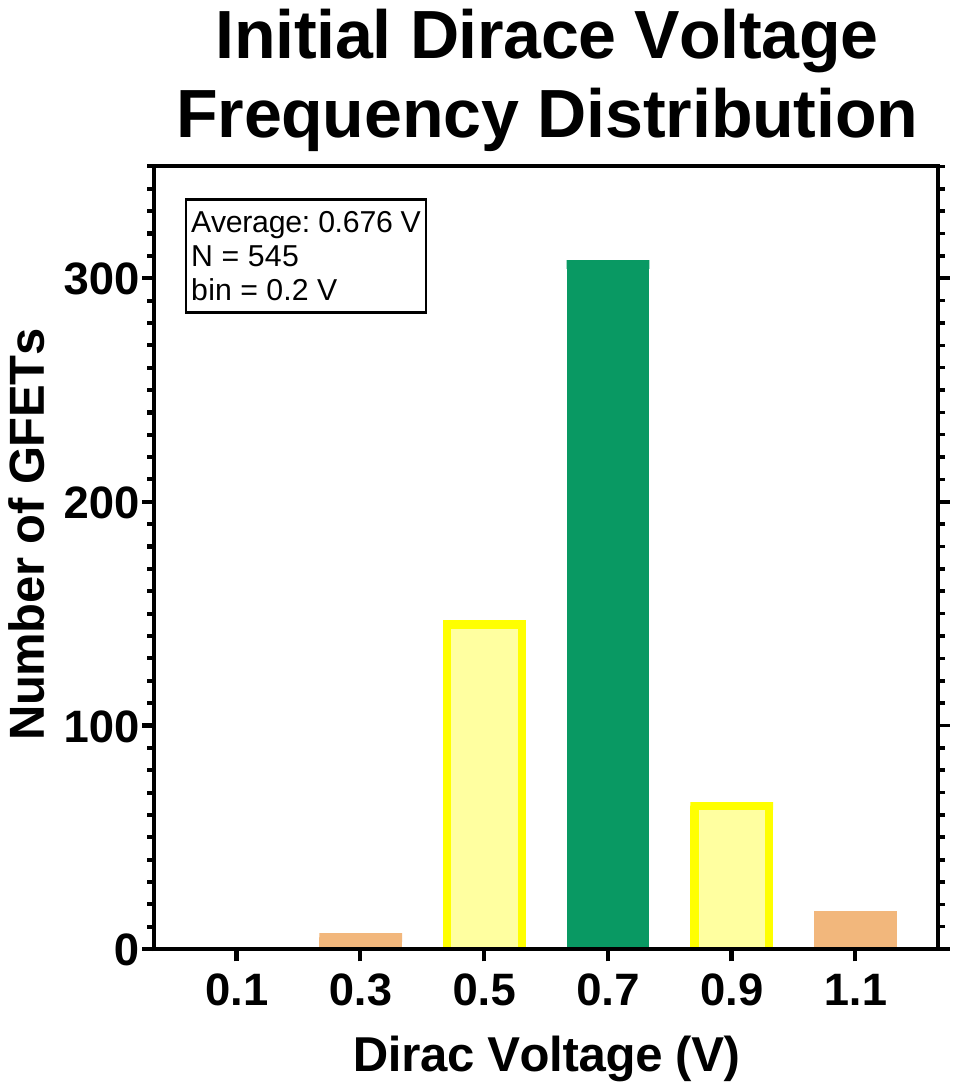

**(a)**

**(b)**

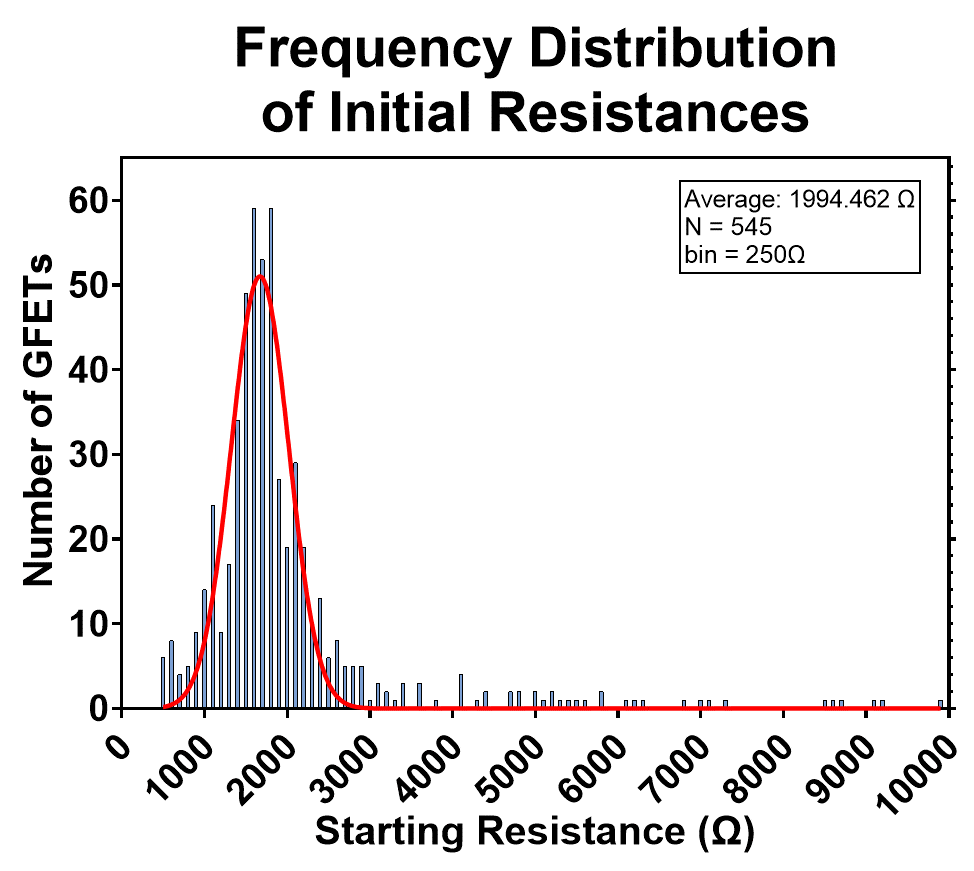

**(c)**

**S4.** Hill’s Equation fitting and LOD calculations.

Hill Fit:

$$V_{D}=\frac{V_{D}^{max}*C^{n}}{K_{D}^{n}+C^{n}}$$

Find Residuals:

$$\sigma=\sqrt{\frac{\sum\left( Hill Fit Point-measured \right)^{2}}{Number of Data Points}}$$

LOD Calculation with $3\sigma$ Analysis:

$$LOD=\frac{3\sigma}{n}$$

**S5.** AFM showing graphene height pre- (left) and post- (right) linker+aptamer attachment. Increase from ~1nm to ~2nm

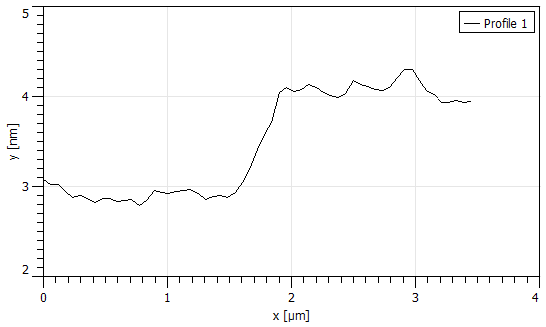

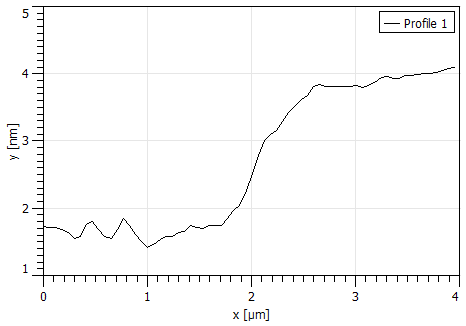

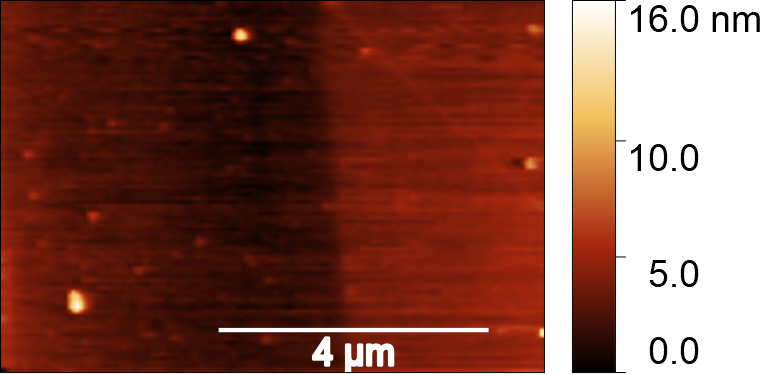

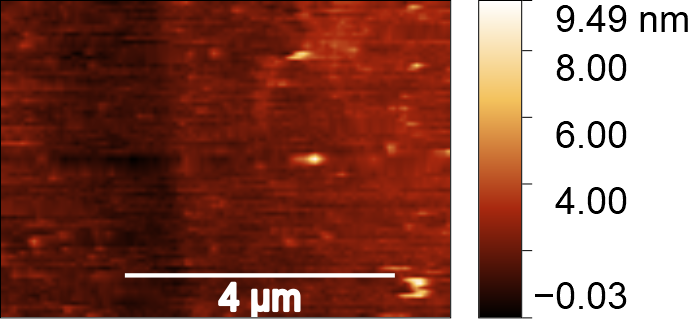

**S6.** Raman Measurements: (top) bare graphene, (middle) linker+aptamer attachment, (bottom) post sensing experiments

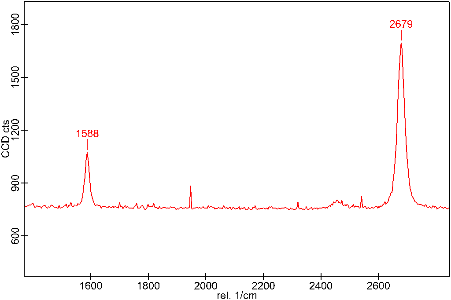

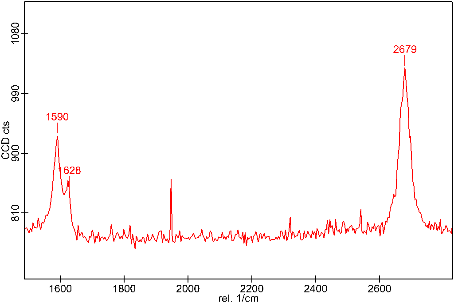

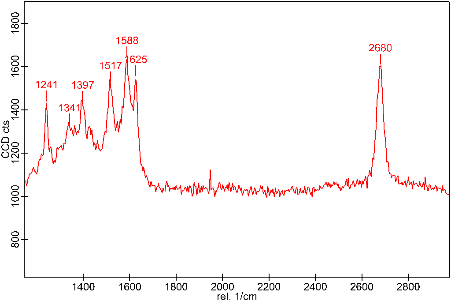

**S7.** *Aptamers*. The aptamers, modified 1C, Universal Aptamer, and H8, were chemical synthesized in a 5’-amine-sequence-3’) configuration by Integrated DNA Technologies (IDT), Coralville, IA. The 1C, 4C, and UA sequences were modified to an unpaired 5’-end nucleotide tail for amine attachment. Upon receipt, the aptamers are diluted to 100 μM concentration using milliQ water following the IDT specification sheet.

1C: CAGCACCGACCTTGTGCTTTGGGAGTGCTGGTCCAAGGGCGTTAATGGACA

The 5’-end was modified to enable linking with PBASE

UA: GGTTTTTACAGCACCACAGACCACCCGCGGATGCCGGTCCCTACGCGTCGCTGTCACGCTGGCTGTTTGTCTTCCTGCC

H8: TAGGGAAGAGAAGGACATATGATAGTGCGGTGAGCCGTCGGACATACAAATACTTGACTAGTACATGACCACTTGA

*Proteins*. The SARS-CoV-2 Spike protein biotinylated (SPN-C82E9), and RSV glycoprotein (RSG-V5221) were bought from ARCO Biosystems while the biotinylated Hemagglutinin (HA) protein (11085-V08H-B) was obtained from Sino Biological.

*Other*. PBASE and PEG were obtained from Sigma-Aldrich. DI water, PDMS, and IPA were obtained from Fisher Scientific.

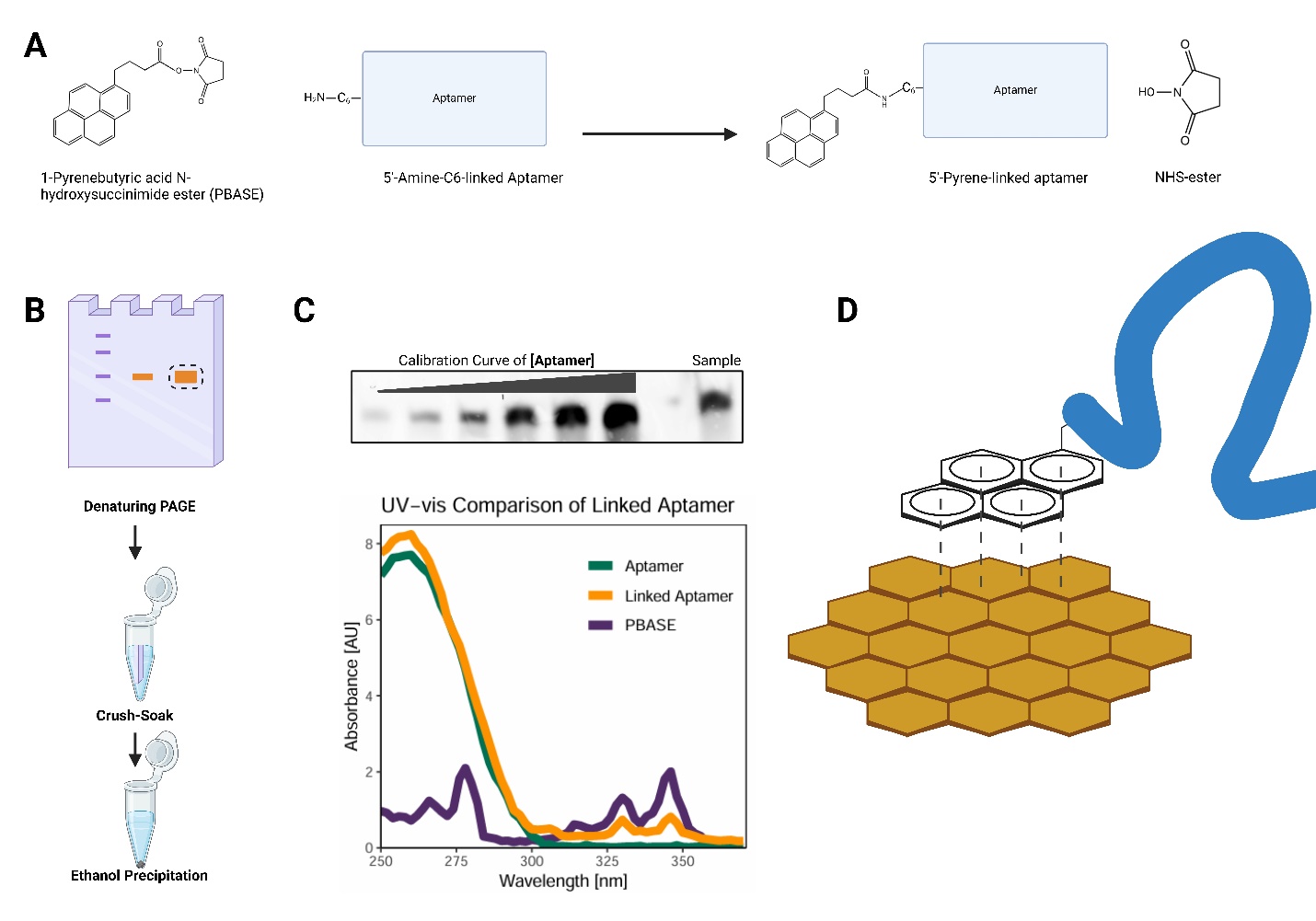

**S8 - Scheme of Pyrene-Aptamer conjugation reaction.**

A) 1-Pyrenebutyric acid N-hydroxysuccinimide (PBASE) conjugation to an aptamer with a 5’ C_6_-amine modification. In slightly alkaline conditions, the NHS ester reacts with the primary amine, linking the pyrene to the aptamer and releasing NHS. B) To purify pyrene-linked aptamer, the reaction is separated via denaturing PAGE and stained with ethidium bromide to visualize DNA using transillumination (365 nm). Linked aptamer is excised and extracted from gel via diffusion and subsequently ethanol precipitated. C) Quality checks for pyrene-linked aptamer include gel densitometry to quantify linked aptamer via a calibration curve (5, 10, 25, 50, 75, 150 pmol of unlinked aptamer visualized ethidium bromide) and UV-vis spectra to confirm PBASE conjugation. The UV-vis spectra of 10 pmol PBASE, 10 pmol linked aptamer, and 10 pmol aptamer are compared. D) Functionalization of graphene. The linked aptamer is then used for concentration-dependent assays of specified targets. Figure created with BioRender.com

Conjugation

The *in vitro* pyrene-aptamer conjugation differs from the previous linking reaction (Kumar et al., 2022) in that it was performed in a microcentrifuge tube instead of on the graphene surface. The goal of this acyl transfer is for the NHS ester group of the 1-Pyrenebutyric acid N-hydroxysuccinimide (PBASE) to interact with the amine group attached to the 5’ end of the aptamer (**S8a**), resulting in a pyrene-linked aptamer that can be attached to the graphene. To achieve this, we adapted a manufacturer’s protocol (<https://www.sigmaaldrich.com/US/en/technical-documents/protocol/genomics/pcr/nhs-ester-oligonucleotide-conjugation>). Briefly, 1 nmol of 5’ NH_2_-C_6_-modified aptamer (IDT) in 10 μL H_2_O was mixed with 10 μL of 1 M NaB buffer, pH 8.5, and 10 μL DMSO for 30 μL total. A 250 nmol excess of PBASE (Sigma Aldrich, Cat No: 457078) (25 μL of a 10 mM solution in DMSO (Sigma Aldrich D8418)), was added to the reaction for a final reaction volume of 55 μL. The reaction was mixed and incubated at 37° C for 2 hours with regular vortexing.

Gel Extraction and Ethanol Precipitation

After the 2-hour incubation reaction, samples were separated via a 6 % polyacrylamide 7.5 M urea gel (National Diagnostics, EC-830) at a constant 35 mA for 90 minutes (**S8b**). After post-staining with ethidium bromide, the gel was imaged using a transilluminator at 365 nm. Linked nucleic acid fragments were excised using a razor, removing excess PBASE and organic solvents. Gel fragments were transferred into a crush soak solution (5 M NaCl, pH 7.5, 1 M Tris·HCl pH 7.5, 0.5 M EDTA pH 8), and pyrene-linked aptamers were extracted via diffusion (5:1 ratio of crush soak buffer volume: gel fragment mass incubated on a rocker for an hour at 37°C). Subsequently, 500 μL of extracted conjugated aptamer in crush soak solution was transferred into a tube containing 1 mL of ice-cold ethanol and 1 μL of glycogen to enable ethanol precipitation. Up to three rounds of crush soak were performed.

Linked aptamers in ethanol were incubated at -20°C before centrifugation at 13,200 rpm for 15 minutes to pellet precipitated linked aptamer. The supernatant was removed, and the pellet was allowed to dry before reconstitution in H_2_O.

Gel Densitometry

The pyrene group of the PBASE interferes with characteristic ssDNA absorbance at 260 nm (Maeda et al., 2001). Therefore, concentration was obtained through quantitative gel densitometry (**S8c**). Using a 6 % polyacrylamide 7.5 M urea gel, a standard curve of known aptamer concentrations and an aliquot of the linked aptamer underwent electrophoresis, ethidium bromide staining, and quantitative imaging [General Electric Typhoon FLA 9500]. A calibration plot of the standard curve band density (raw volume) vs. concentration was used to calculate the concentration of the linked aptamer. The pyrene-conjugated aptamer was then aliquoted to 50 μL of 10 μM for downstream applications.

UV-vis Spectrophotometry

PBASE has characteristic UV adsorption peaks at 325 nm and 341 nm (Bao et al., 2010) absent in aptamers. Therefore, comparing UV-vis peaks from PBASE, the unlinked aptamer, and a sample of linked aptamer ensures a successful linking reaction (**S8c**).

Bao, Q., Zhang, H., Yang, J. X., Wang, S., Tang, D. Y., Jose, R., Ramakrishna, S., Lim, C. T., & Loh, K. P. (2010). Graphene–Polymer Nanofiber Membrane for Ultrafast Photonics. *Advanced Functional Materials*, *20*(5), 782–791. https://doi.org/10.1002/ADFM.200901658

Kumar, N., Rana, M., Geiwitz, M., Khan, N. I., Catalano, M., Ortiz-Marquez, J. C., Kitadai, H., Weber, A., Dweik, B., Ling, X., van Opijnen, T., Argun, A. A., & Burch, K. S. (2022). Rapid, Multianalyte Detection of Opioid Metabolites in Wastewater. *ACS Nano*, *16*(3), 3704–3714. https://doi.org/10.1021/acsnano.1c07094

Maeda, H., Inoue, Y., Ishida, H., & Mizuno, K. (2001). UV Absorption and Fluorescence Properties of Pyrene Derivatives Having Trimethylsilyl, Trimethylgermyl, and Trimethylstannyl Groups. *Chemistry Letters*, *30*(12), 1224–1225. https://doi.org/10.1246/CL.2001.1224
